# Surveillance strategies for the detection of new SARS-CoV-2 variants across epidemiological contexts

**DOI:** 10.1101/2023.05.09.23289744

**Authors:** Kirstin I. Oliveira Roster, Stephen M. Kissler, Enoma Omoregie, Jade C. Wang, Helly Amin, Steve Di Lonardo, Scott Hughes, Yonatan H. Grad

## Abstract

Rapid identification of new SARS-CoV-2 variants is a critical component of the public health response to the COVID-19 pandemic. However, we lack a quantitative framework to assess the expected performance of sampling strategies in varying epidemic contexts. To address this gap, we used a multi-patch stochastic model of SARS-CoV-2 spread in New York City to evaluate the impact of the volume of testing and sequencing, geographic representativeness of sampling, location and timing of variant emergence, and relative variant transmissibility on the time to first detection of a new variant. The strategy of targeted sampling of likely emergence locations offered the most improvement in detection speed. Increasing sequencing capacity reduced detection time more than increasing testing volumes. The relative transmissibility of the new variant and the epidemic context of variant emergence also influenced detection times, showing that individual surveillance strategies can result in a wide range of detection outcomes, depending on the underlying dynamics of the circulating variants. These findings help contextualize the design, interpretation, and trade-offs of genomic surveillance strategies.

## Introduction

Genomic surveillance is an important tool for public health response to infectious disease (1,2). For pathogens such as SARS-CoV-2, genomic surveillance has enabled rapid identification and characterization of genetic variants that differ in transmissibility, virulence, and antigenic space, and thus informed disease control policies and the design and clinical use of therapeutics and vaccines (3–7).

Many questions remain about the impact of sampling strategies on the time to first detection of a variant. Guidelines suggest specific sequencing rates or fixed sequencing volumes (7)(8), depending on available resources, logistical considerations, overall SARS-CoV-2 prevalence, and surveillance objectives. They also emphasize the importance of sequencing cases that accurately reflect all SARS-CoV-2 infections, which requires representativeness of both testing and sequencing. In low- and middle-income countries, increasing testing volume as a way of improving the representativeness of sampling is most important to reducing variant detection times (9). However, we lack a general quantitative assessment of sampling strategies to detect new variants—where ‘new’ refers to local emergence of a novel variant or the importation of an emerging, known or an unknown variant—within a target detection time that considers the role of geographic representativeness of sampling, the optimal testing and sequencing volumes, and epidemic context.

To address this gap, we simulated the impact of a set of surveillance strategies on variant detection across a range of contexts. We developed a multi-patch stochastic transmission model for SARS-CoV-2 in New York City (NYC) and incorporated empirical human mobility data for the geographic dispersal of pathogens. We chose NYC as a case study given its experience with genomic surveillance and publicly available data on testing, sequencing, and mobility (10,11). We simulated the introduction of new variants with varying levels of transmissibility in locations across the city and at multiple introduction times relative to the introduction of the previously dominant variant. We then simulated testing and sequencing scenarios, varying both the volume and distribution of sampling, and computed the time to first detection, the overall burden of disease, and the geographic variability of the disease burden under each strategy. By developing this framework, we aimed to contextualize decision-making on genomic surveillance within the diversity of possible disease scenarios.

## Methods

### Data

Baseline COVID-19 testing rates (609 tests per 100,000 residents per week) and sequencing rates for NYC were obtained from the NYC Department of Health and Mental Hygiene (NYC DOHMH) (11) from December 2020 until November 2021 at the geographic resolution of modified ZIP-code tabulation areas (MODZCTAs). Mobility data were obtained from Meta *via* the Facebook Data for Good Initiative (12), which reported the physical locations of anonymized app users within 600m-by-600m tiles in 8-hour intervals. These data were aggregated to both MODZCTAs and boroughs and used to construct a mixing matrix estimating the rate of interpersonal encounters among the residents of NYC. We used data from the United States Census Bureau to define mappings between MODZCTAs, tiles, and boroughs. The main analysis was conducted at the geographic scale of boroughs. We conducted a sensitivity analysis at the level of MODZCTAs. Full details are provided in the **Supplementary Materials and Methods**.

### Model structure

To simulate the introduction and subsequent transmission of a novel SARS-CoV-2 variant, we constructed a multi-patch, two-variant stochastic compartmental model that builds on the basic structure of a Susceptible-Exposed-Infectious-Recovered-Susceptible (SEIRS) model. A first variant was seeded in the population and a single index case of a novel variant was introduced at varying times and in varying locations. The new variant was simulated to be more infectious than the previously circulating variant. In sensitivity analyses (see Supplemental Material), we considered variants that also had greater and faster immune evasion. Individuals progressed stochastically through the states defined by variant characteristics (cross-protection, transmission probability, duration of latent and infectious periods) and the number of individuals in each compartment. The probability of transmission between geographic locations (patches representing a borough or MODZCTA) was governed by the contact between boroughs (as observed through human mobility), and the relative quantities of infectious and susceptible residents. We accounted for imperfect test sensitivity and specificity and for infection-induced behavioral changes, such as reducing contacts in response to a positive test. We then assessed the impact on detection outcomes of different characteristics of the surveillance strategy and the epidemiological context, specifically the volume of testing and sequencing, the geographic distribution of testing, the introduction time and location of the second variant relative to the first, the probability of transmission, and the connectivity (inward and outward mobility) of introduction and sampling locations. Full details on the model structure are provided in the **Supplementary Materials and Methods**. Code is available at github.com/gradlab/detectingsarscov2-variants.

### Statistical analysis

The main outcomes in this study were the time to variant detection (the number of days between when the index case becomes infectious and laboratory confirmation of the new variant among sequenced specimens), the cumulative number of infections, and the variation in cumulative infections across locations. We ran 100 simulations per scenario and calculated the arithmetic means, medians, and confidence intervals of the main outcomes across simulations. For sampling schemes, we considered (1) the distribution of test volume in New York City provided to DOHMH, which we termed ‘baseline’ testing, (2) test volumes distributed by population density, and (3) test volumes distributed randomly across locations. We also considered “focused testing scenarios” in which 20-100% of tests were allocated in a single location, with the remaining tests distributed evenly among the other locations according to their population size. For total testing volumes, we considered a range between 5% and 300% of the reported testing volume. For sequencing rates, we considered a range between 1% and 90% of all positive tests to be selected for sequencing. In a sensitivity analysis, we evaluated fixed sequencing quantities instead of sequencing proportions, which distinguished the marginal contribution of sequencing versus testing alone.

When all introduction times produced qualitatively similar results, we reported in the main text the results from the scenario in which the new variant is introduced just before the outbreak peak of the previously dominating variant (at *t* = 50 days). In this scenario, detection was expected to be at or near its slowest, due to high background prevalence and depletion of susceptible individuals. Full results for all parameter combinations are provided in the **Supplementary Data**.

## Results

### Geographic sampling strategy

Relative to the baseline volume and distribution of testing and sequencing in NYC (the “baseline” testing and sequencing strategy), detection times were similarly distributed when test volumes were allocated to be (a) proportional to the population density or (b) uniformly at random across locations (**Fig. 1**). This similarity across geographic sampling strategies was unaffected by the outcome measure used as well as the timing and location of the new variant’s introduction. However, the geographic sampling strategy affected detection outcomes if the introduction location of the new variant was oversampled. Allocating a greater proportion of tests in a single location reduced detection times and cumulative infections of variants emerging in that location but increased detection times of variants that first appeared elsewhere (**Fig. 2**). This effect was especially pronounced in Staten Island and Brooklyn, but weaker in Manhattan (**Supplementary Fig. S5**).

**Figure 1.**
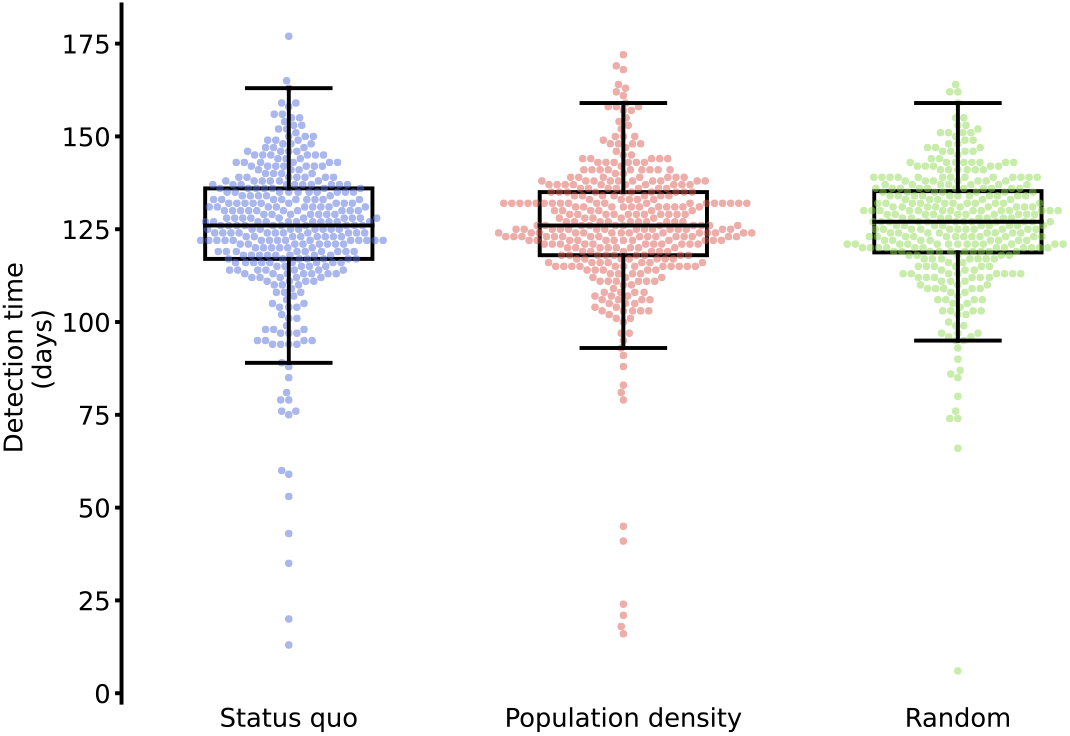
Distribution of detection times by geographic sampling strategy. Points depict the time between variant introduction and detection in days for the scenarios where tests are sampled geographically according to the baseline testing strategy, proportionally to population size, or randomly across New York City (at variant introduction 50 days after the prior variant, 30% of baseline test volume, and 10% sequencing rate). Boxes and whiskers depict the minimum, lower 25%, median, upper 75%, and maximum detection times.

**Figure 2.**
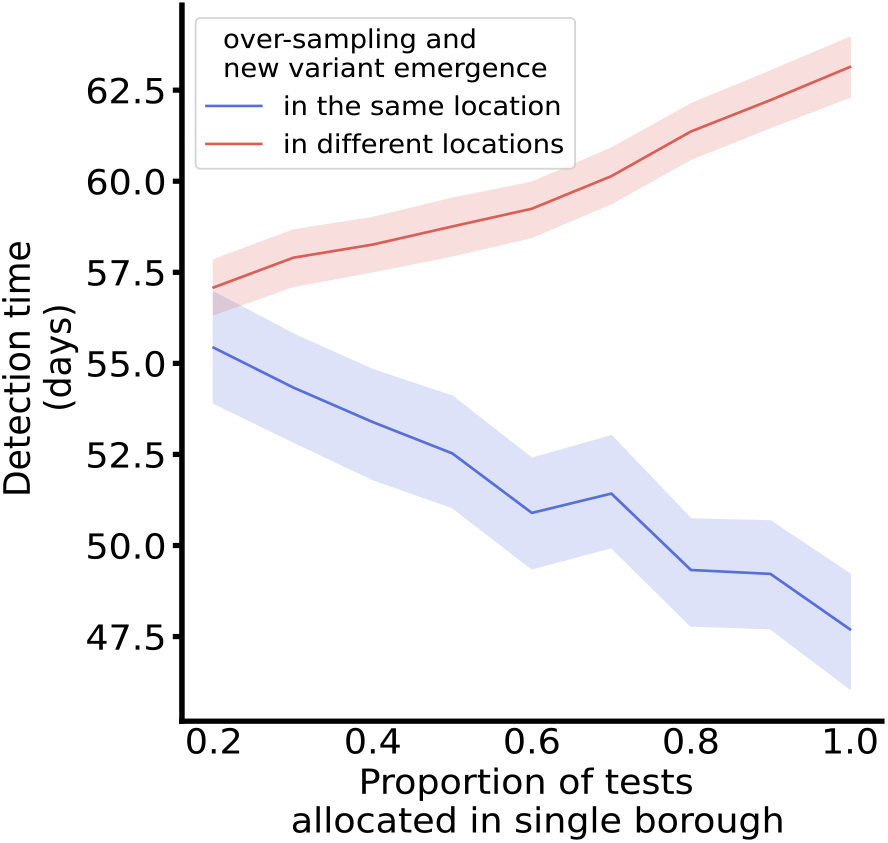
Detection time by proportion of tests allocated in a single location. Lines depict the average detection time for scenarios where between 20% and 100% of tests are sampled from a single location, and the remaining tests are evenly distributed across the remaining locations by population size. The lines distinguish between scenarios where the variant emerged in the primary allocation location, i.e., test over-sampling and emergence occurred in the same location (blue), and scenarios where the variant emerged in one of the other locations, i.e., test over-sampling and emergence occurred in different locations (red). Ribbons depict the 95% confidence interval for the detection time.

### Testing and sequencing volumes

Outcomes varied considerably across testing and sequencing rates, with higher rates leading to faster detection, fewer cases, and less variation in cumulative infections across locations (**Fig. 3**). In accordance with sampling guidelines for well-resourced settings (7), we assumed that a fixed percentage of tests was sequenced. Thus, increasing the number of tests also increased the number of sequenced samples.

**Figure 3.**
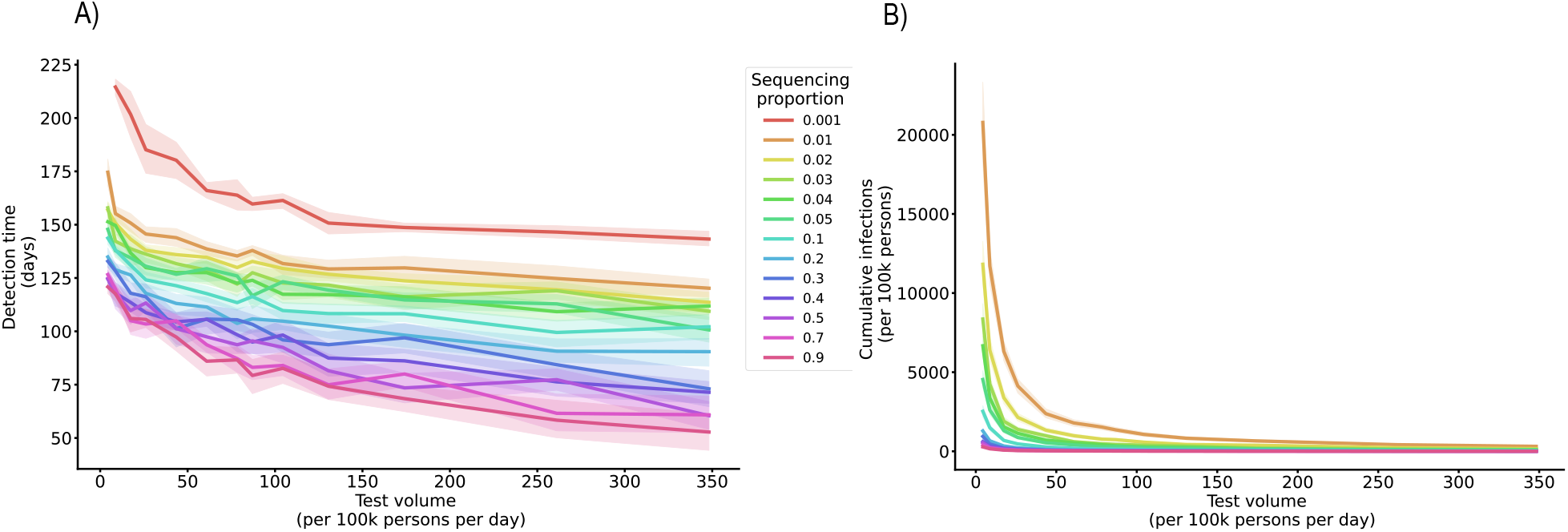
Detection outcomes by test quantity and sequencing rate. Lines depict the mean duration between variant introduction and detection in days (A) and the cumulative infections upon detection (B) as a function of daily testing volume (given new variant introduction 50 days after the prior variant, baseline test strategy). Ribbons depict the 95% confidence interval for the detection time. Colors represent proportions of tests selected for sequencing.

To better understand the individual contributions of testing and sequencing, we fixed the quantities of samples selected for sequencing at varying testing volumes. Fixed sequencing volumes were implemented as a cap on the maximum number of samples that can be sequenced per day. The actual number of sequenced cases depended on the test positivity rate.

The improvement in variant detection with increasing test volumes at a given sequencing proportion was driven by the increase in sequencing volume rather than test volume. At all levels of testing, increasing the number of sequenced samples reduced the detection time, while increasing testing alone had little impact on new variant detection (**Fig. 4**).

**Figure 4.**
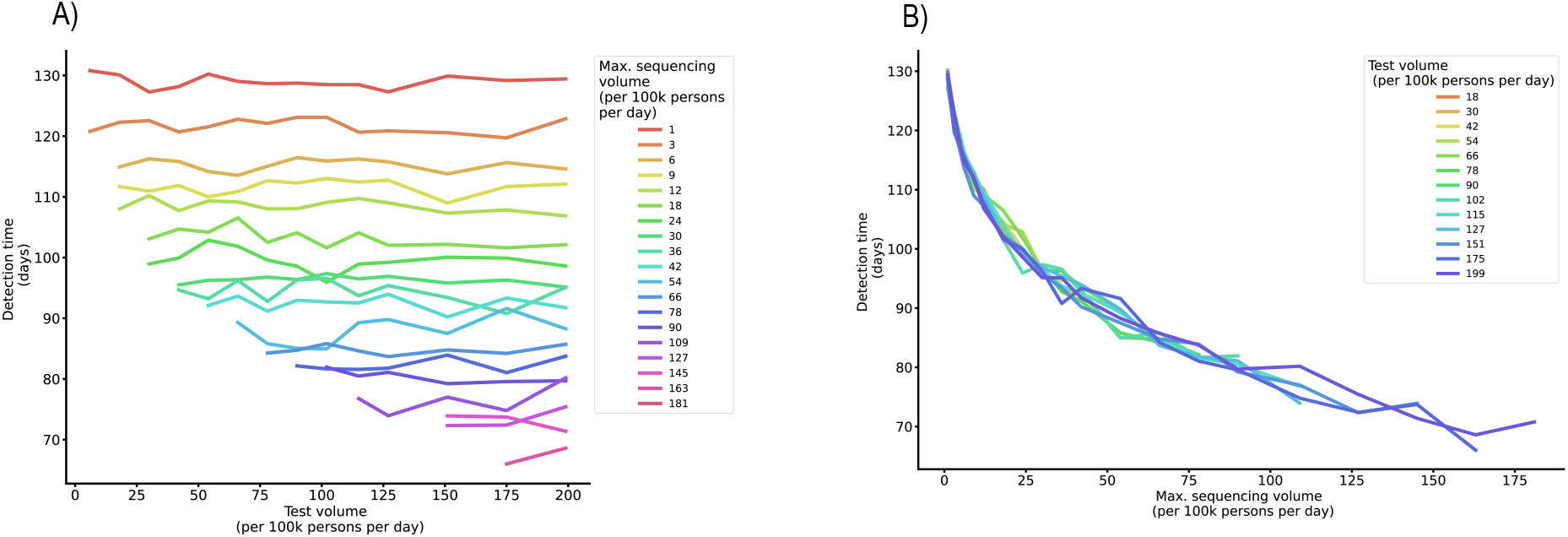
Detection time by test volume and fixed sequencing capacity. Lines depict the mean duration between variant introduction and detection in days as a function of daily testing volume, colored by the maximum sequencing volume (A), and as a function of daily maximum sequencing volume, colored by the test volume (B) (at variant introduction 50 days after the prior variant and baseline sampling strategy).

We also considered an alternative interpretation of the sequencing cap, where the sequencing volume depended on both the test volume and the positivity rate (**Supplement 3; Supplementary Materials and Methods**). The results from this sensitivity analysis fall between the fixed volume and fixed rate analyses (**Figs. 3 and 4**). Raising testing capacity improved detection times for low levels of testing (up to 50-75 tests per 100k persons). At higher levels of testing, improvements in detection time were driven primarily by increased sequencing capacity (**Supplementary Fig. 3**).

### Emergence context

We compared introduction times of the new variant as an approximation for varying background prevalence of the previously circulating variant and the population susceptibility to infection.

When the second, more transmissible variant was introduced into a fully susceptible population together with the first variant (at *t* = 0), the second variant was more likely to dominate due to its increased transmissibility. Under this scenario, the extinction probability of the second variant (defined as the likelihood that a variant will cause no more than 10 infections) was only 9.6% under the baseline sampling strategy. Both variants generally persisted through the duration of the simulation, though the second variant caused more infections. Consequently, at a *t=0* introduction time, the second variant was detected in under 33 days in 95% of simulations. If the second variant was introduced after the peak of the first variant’s outbreak (at *t* = 80 or *t* = 100), the second variant had a high probability of extinction (64.6 and 84.2%, respectively), and if it persisted, it was detected later (at least 56 and 37 days after introduction in 95% of simulations, respectively). The greatest range of disease dynamics and consequently detection times was observed when the second variant was introduced just before the peak of the first variant (at *t* = 50), with detection times ranging from 16 to 145 days (**Fig 5**).

**Figure 5.**
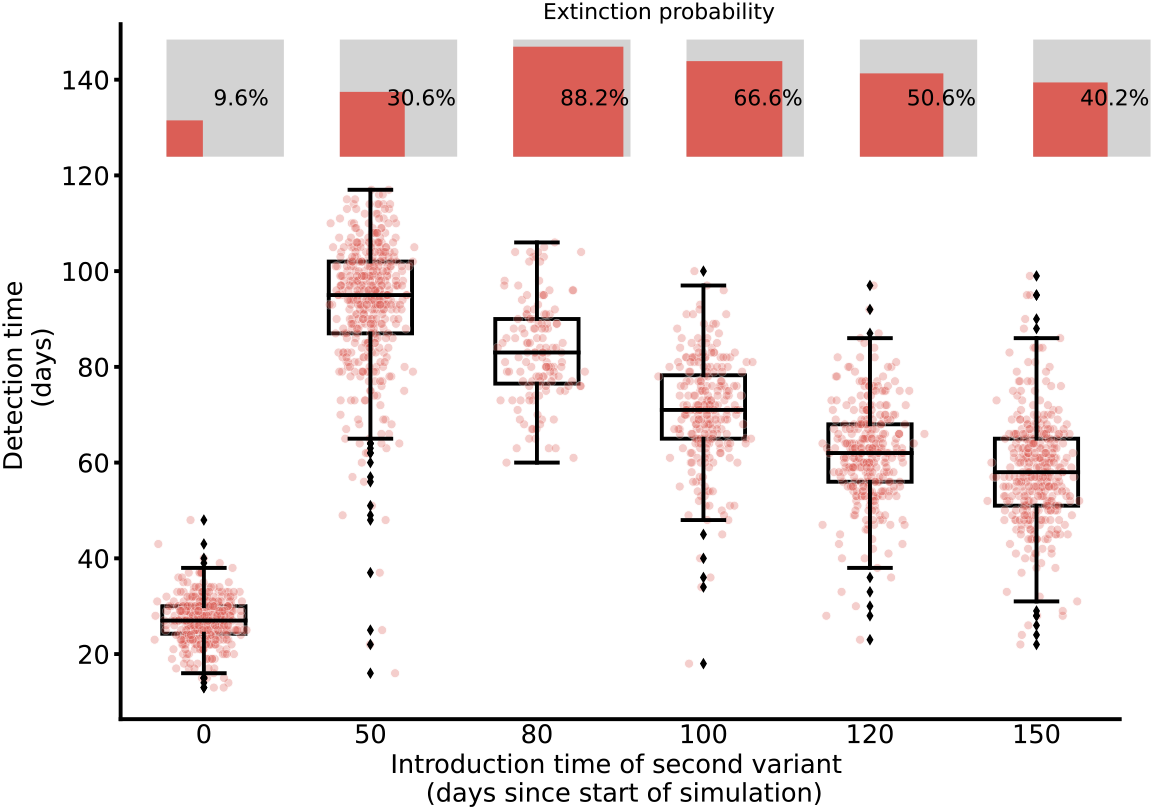
Detection time of a novel variant across introduction times. Points depict the time between variant introduction and detection in days for different introduction times (with baseline distribution of tests, 30% of baseline test quantity, and sequencing rate 10%). Points are jittered horizontally to help visualize the distribution. Boxes and whiskers depict the minimum, lower 25%, median, upper 75%, and maximum detection times. The extinction probability for each scenario is depicted using inset squares, where the relative area of the red square is proportional to the extinction probability.

The introduction location did not significantly impact the detection time or cumulative disease burden across the city but did influence where infections occurred .The number of infections was highest in locations with the highest mobility connectivity to the emergence location, which was either the introduction location itself or other locations, depending on the mobility matrix. Emergence in Staten Island, for example, produced infections primarily within Staten Island, while emergence in Manhattan led to a high number of infections in Brooklyn and Queens (**Supplementary Fig. S4**).

### Variant characteristics

We compared variants with different levels of transmissibility, varying the probability of infection given an infectious contact from *β* = 0.21 to *β* = 0.5 (contrasting with the transmissibility of the first variant of *β* = 0.2). This transmission parameter affected the disease dynamics, with more transmissible variants spreading more quickly, leading to earlier detection. All transmission rates yielded a wide range of cumulative infections at detection time (**Fig. 6**).

**Figure 6.**
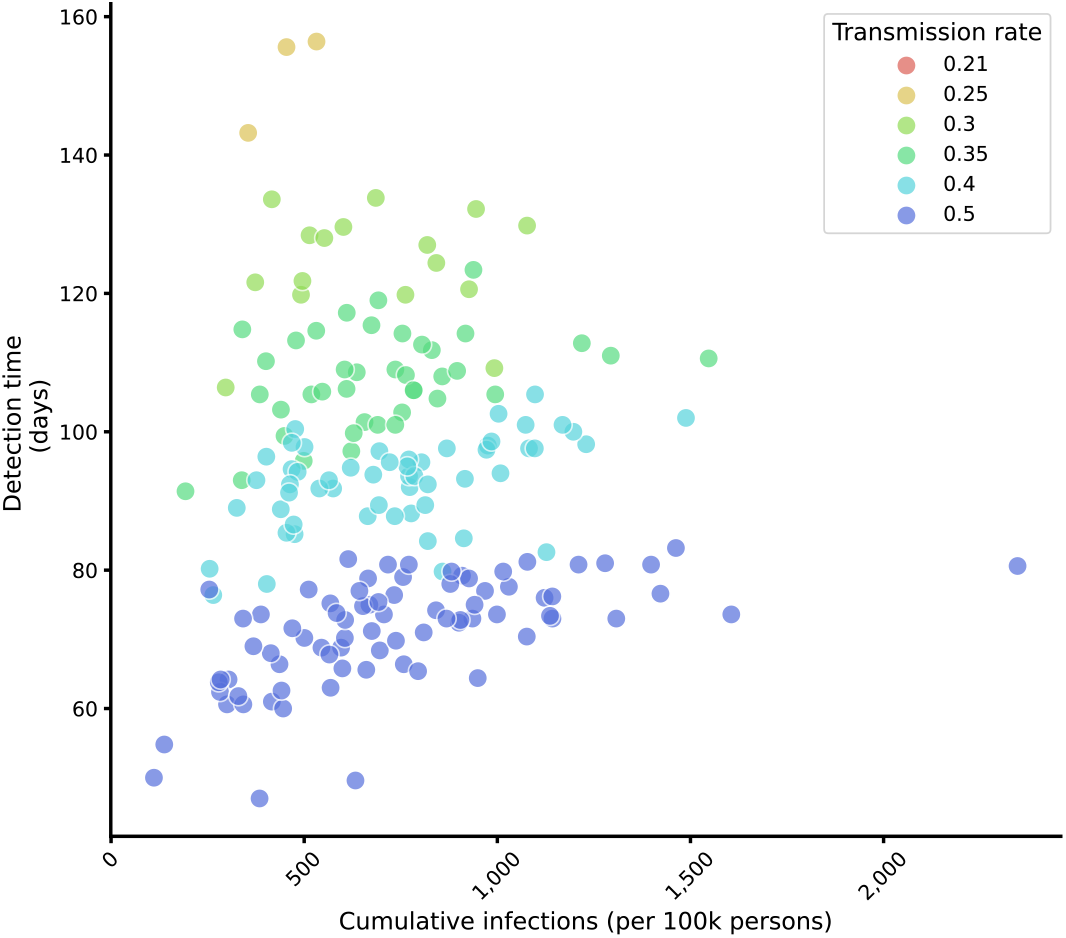
Detection time by cumulative infections for different transmission rates. Points depict the mean detection time and cumulative number of infections upon detection, averaged across 100 simulations of each introduction location, for each of the six transmission probabilities, represented by different colors (at variant introduction 50 days after the prior variant, baseline distribution of tests, 30% of baseline test quantity, and sequencing rate 10%). The baseline transmission rate of the pre-existing variant is β = 0.2.

## Discussion

This study provides an assessment of testing and sequencing strategies for the detection of new SARS-CoV-2 variants to help inform genomic surveillance policies. We considered varying quantities and distributions of resources within a wide range of potential settings for variant emergence and assessed how they influenced variant detection times and the undetected disease burden.

Our results confirm that variant detection is governed by both the surveillance strategy and the epidemic dynamics in which the new variant arises (13). The relative transmissibility of the new variant as well as the context of variant emergence influenced its speed of spread and extinction probability, which in turn affected detection outcomes (Figs. 5 and 6).

Surveillance guidelines stipulate the need for representative sampling strategies, which require not only that a random subset of positive cases be chosen for sequencing, but that positive cases also accurately reflect infections across the population; non-representative sampling delays the detection of new variants (14). Nonrepresentative testing and sequencing must be considered separately and jointly, along dimensions such as demography, socioeconomic factors, disease outcome, and geography. In this study, we assessed the role of geography in representative sampling. For variation in the geographic distribution of sequencing, other work found that if sampling was not representative, then raising test volume improved variant detection times more than raising sequencing rates (9). Our study considered the geographic distribution of testing, as well as the volume of both testing and sequencing, and found that in the context of random sampling of tests for sequencing, improvements in detection time were driven primarily by increases in sequencing volume rather than testing volume (**Fig. 4**). The geographic distribution of residents receiving tests impacted detection outcomes only via the proximity to the emergence location of the new variant (**Fig. 2**), though careful test distribution matters to many related public health and social equity objectives (14–16). Oversampling emergence locations improved detection outcomes, underscoring the importance of targeted sequencing, for example at ports of entry or of patients with prolonged viral replication (8). The connectivity of the introduction location did not impact detection times but did affect where infections occurred before variant detection (**Supplement 3**). Variants that emerged among residents of boroughs with more inward and outward mobility produced more infections in other boroughs. In our simulations, a variant first appearing in a resident of Manhattan, for example, caused more infections on average in Brooklyn and Queens than in Manhattan itself. Failing to adequately sample locations near emergence or those highly connected to emergence locations will lead to a disproportionate number of infections in those locations.

The number of undetected infections varied widely for a given transmission rate, even at fixed detection times (**Fig 6**). This result demonstrated the challenge of understanding the epidemiologic scenario on discovery of a new variant and the need for combining pathogen genome sequencing with other forms of surveillance. More work is also needed to understand whether optimal surveillance strategies differ if the primary objective is monitoring or detecting variants and how to position genomic surveillance within the broader landscape of sometimes competing public health objectives.

The model in this study was designed to be simple, while accounting for the most important factors affecting testing and sequencing, and to help attain a qualitative understanding of which parameters influence detection times and undetected infections. Consequently, the simulation results, such as the detection times, should not be interpreted as predictions. Specific simplifications include the modeling of single introductions of a novel variant, rather than accounting for multiple introductions or several variants. We also assumed homogeneous mixing within locations and did not account for age structure, social networks, or social determinants of health. SARS-CoV-2 infection risk varies across socioeconomic and demographic groups, due in part to variability in the average number of contacts, vaccine uptake, longand shortdistance mobility, comorbidities linked to more severe disease outcomes, and other social factors (17–19). While we incorporated neighborhood-level variations in movement, we did not include within-neighborhood heterogeneity or between-neighborhood variation in social determinants of health. A basic first analysis suggests that household income correlates negatively with undetected infections in our simulations (**Supplement 4**). Future work may explore how these heterogeneities influence emergence locations of new variants, disease dynamics, and consequently detection outcomes.

The model in this study also took a simplified perspective of genomic surveillance processes.

We assumed random sampling of positive tests and did not account for variations in specimen quality across testing sites or in access to testing, which may cloud estimates of the prevalence of circulating variants (20). In this sense, our model takes an idealized view of our capacity to sample randomly from the population in each borough or zip code.

Even though we incorporated human mobility data, the model did not account for human responses to the epidemiological situation, such as reduced contact rates during periods of high or rising prevalence, or changes in public health policies like mask mandates. Contact patterns vary over time, not just across places, and future studies should examine how new variant detection changes when incorporating time-varying human behavior.

Detection of novel variants remains a critical component of the response to the COVID-19 pandemic. Emerging empirical evidence on genomic surveillance of SARS-CoV-2 variants has allowed public health agencies to provide guidance on sampling strategies to detect and monitor variants, though more research is needed to anticipate the impact of these strategies under as yet unseen epidemiologic settings. This modeling study aimed to contribute to these ongoing efforts to assess variant detection strategies, by simulating detection outcomes for varying testing and sequencing rates in NYC. We focused on the role of geography in representative sampling as well as the context of variant emergence. The epidemiologic context, including emergence timing and background prevalence, played an important role in shaping detection times and undetected disease burdens. Targeted sampling of emergence locations was the primary aspect of geographical representativeness examined in our model that improved detection outcomes. Increased testing is an important tool to enable more representative sampling (9), though in well-resourced settings with random sampling, as was assumed in this case study of NYC, increasing sequencing capacity rather than testing had a larger impact on improving detection speeds.

## Data Availability

Code and data are available at github.com/gradlab/detecting-sarscov2-variants.

## Acknowledgments

The authors thank Faten Takai for helpful feedback on the manuscript and the Public Health Lab whole genome sequencing and data units.

## Supplementary Materials and Methods

### Data

#### Mobility

Through its Facebook Data for Good initiative, Meta provides aggregated and anonymized movement data from users who have enabled location sharing (12). It is available in 8-hour intervals and with a maximum geographic resolution of 600-by-600m tile sizes. We leveraged this data to compute the average share of users moving between geographic tiles on a given day, aggregated to both modified zip code areas (MODZCTA) and boroughs. This percentage was rescaled to the population size of each location, under the assumption that Facebook users who activate location sharing are representative of NYC residents, which may not be true in practice and may limit our ability to accurately capture human movement across the city. The movements were captured in the raw mobility matrix *M*, where an entry *M*_{*i*−>*j*}_ indicates the number of movements from location *i* to location *j*. We then computed a contact matrix from the mobility matrix under the assumption of homogeneous mixing. The probability that a resident of location *i* has contact with a resident of location *j* in any location *k* was defined as:

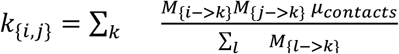

Where *μ*_*cantacts*_ is the average number of contacts per person, *M*_{*i*−>*k*}_ is the number of individuals moving from location *i* to location *k*, and Σ_*l*_ *M*_{*l*−>*k*}_ is the sum of all individuals moving to location *k*. This contact matrix determined the coupling strength of two locations in the mathematical model, and therefore influenced the likelihood for an infection to spread between the two locations.

*Testing and Sequencing Rates*

Weekly COVID-19 testing rates and sequencing rates by MODZCTA are published by the NYC Department of Health and Mental Hygiene (16). The baseline test rate was calculated as the daily average from December 2020 to November 2021.

### Compartmental Model Structure

We implemented a multi-patch, two-variant stochastic compartmental model which builds upon the basic structure of an SEIRS model. Exposure, testing, sequencing, isolation, recovery, waning (cross-)protection, and reinfection were treated as stochastic events. The model explicitly incorporated human mobility, testing and sequencing rates, as well as test sensitivity and specificity. It also included control measures to account for the fact that testing strategies may impact disease dynamics if infectious individuals reduce their contacts upon receiving a positive test result.

The model structure is presented in Supplemental Figure 1 and described in more detail below. The corresponding parameters are listed in Supplemental Table 1. Susceptible (*S*) individuals may be exposed (*E*) to one of two variants, represented by subscripts 1 and 2, or they may isolate if they receive a false positive test result (*S*_*q*_). After a latent period, an exposed individual moves to one of five infectious compartments depending on their reporting and isolation outcome: an infection may be unreported (*I*_*U*_), for example if it is asymptomatic or detected only via an at-home test. A positive case may be detected at an official test center (*I*_*T*_), leading the person to isolate (*I*_*Tq*_) or not (*I*_*Tnq*_). Finally, the positive test may also be sequenced (*I*_*G*_), again separating individuals by their isolation status (*I*_*Gq*_, *I*_*Gnq*_). After recovery, individuals have temporary full protection against reinfection with either variant while they remain in compartments *R*_*U*_, *R*_*T*_, *R*_*G*_. Full protection wanes over time at variant-specific rates, making individuals susceptible to reinfection with one (*S*_1{1}_, *S*_2{1}_, *S*_1{2}_, *S*_2{2}_) or both variants (*S*_12{1}_, *S*_12{2}_). The model incorporates a variant-specific leaky immunity parameter (*a*_11_, *a*_12_, *a*_21_, *a*_22_), which defines the reduction in the probability of reinfection after full protection has waned.

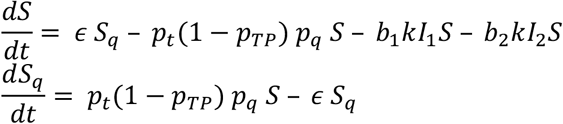

*Variant 1:*

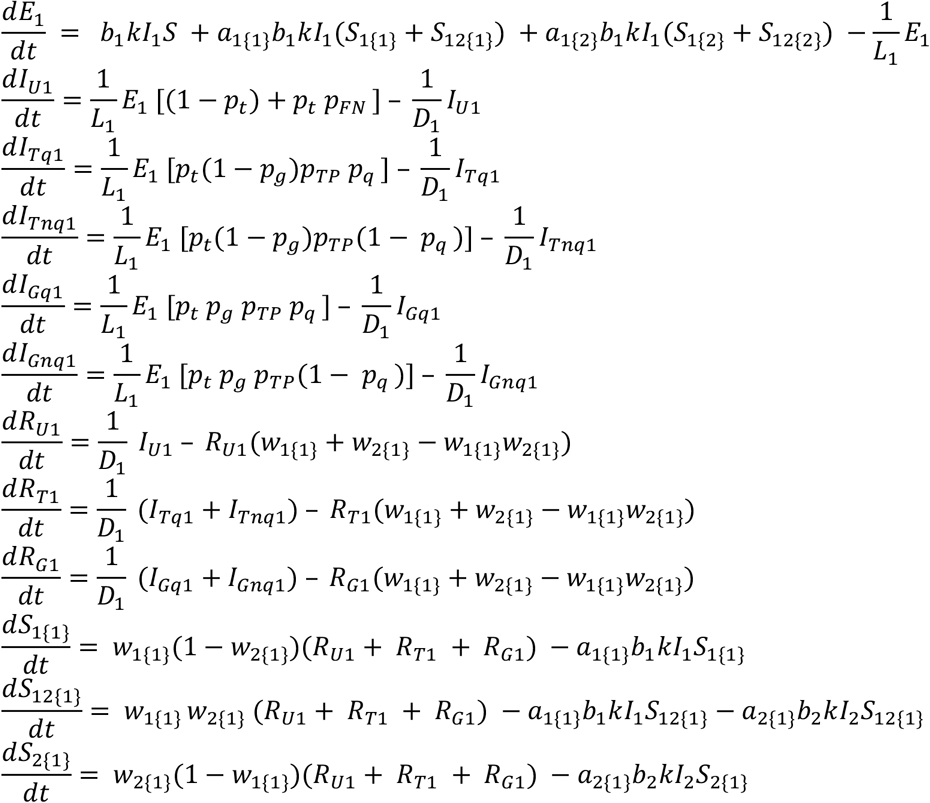

*Variant 2:*

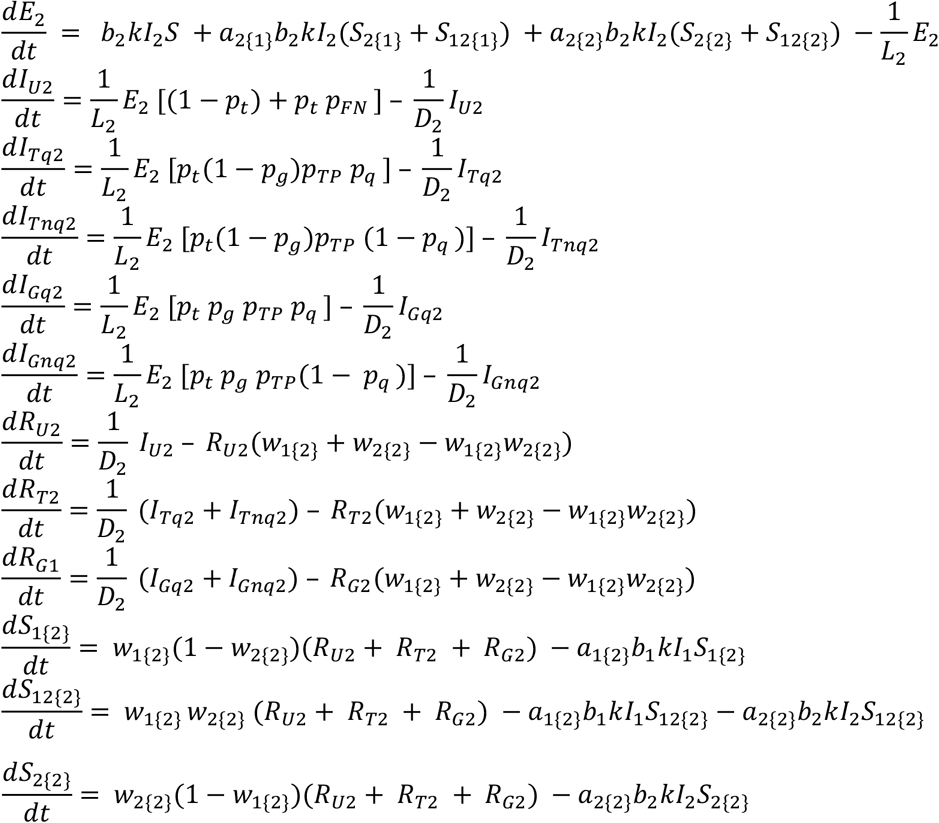

where *I*_1_ = *I*_*U*1_ + *I*_*Tq*1_ + *I*_*Tqn*1_ + *I*_*Gq*1_ + *I*_*Gnq*1_ and *I*_2_ = *I*_*U*2_ + *I*_*Tq*2_ + *I*_*Tnq*2_ + *I*_*Gq*2_ + *I*_*Gnq*2_

**Supplemental Figure S1.**
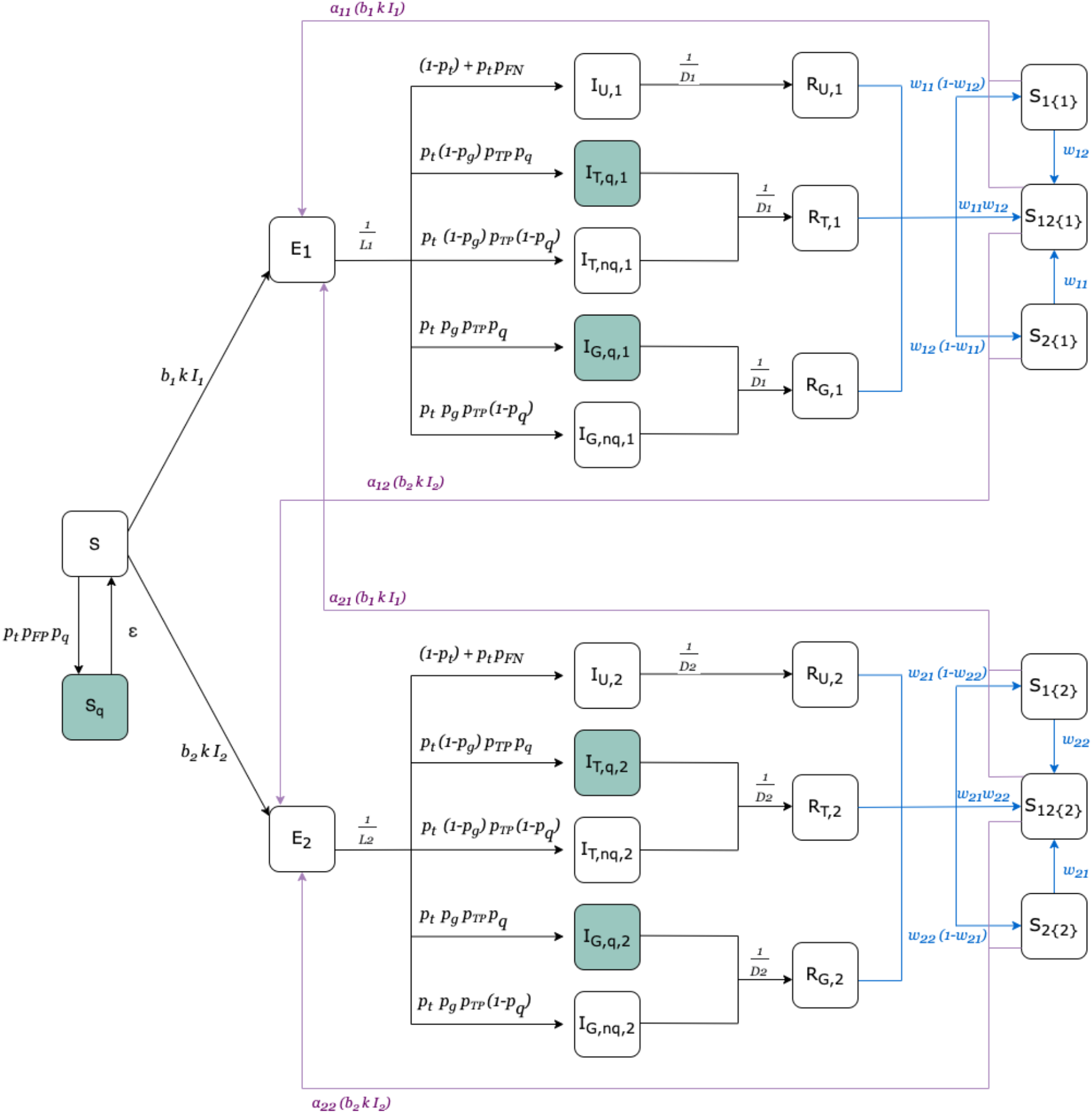
Model structure.

Where *p*_t_ is the test rate, *p*_g_ is the sequencing rate, *p*_*q*_ is the control measure compliance rate, *p*_6:_ is the true positive rate, *a*_*x*{*y*}_ is the leaky immunity parameter, *w*_*x*{*y*}_ is the waning full immunity parameter, *b* is the probability of infection given contact, *L* is the average duration of the latent period, *D* is the average duration of the infectious period, *k* is the contact matrix, and *ϵ* is the rate of ending control measures if false positive.

**Table S1.** Parameters

| Parameter | Description |
| --- | --- |
| $p_t$ | Test rate (share of population) |
| $p_g$ | Sequencing rate (share of positive tests) |
| $p_q$ | Control measure compliance rate |
| $p_{TP}$ | True positive rate of test instrument |
| $a_{x\{y\}}$ | Leaky immunity parameter: percent susceptibility to infection with variant $x$ after infection with variant $y$ |
| $w_{x\{y\}}$ | Waning full immunity parameter: rate of loss of immunity against variant $x$ after infection with variant $y$ |
| $b$ | Probability of infection given a contact with an infectious individual |
| $L$ | Average duration of latent period |
| $D$ | Average duration of infectious period |
| $k$ | Contact matrix |
| $\epsilon$ | Rate of ending control measures with false positive test result |

### Simulations

#### Geographic Test Distribution

We implement the following primary strategies for testing allocation. Supplemental Figure 2 shows an example of the test rates under each scenario.

- Baseline: Tests are allocated according to the current NYC strategy, computed as the average allocation and daily test quantity between December 2020 and November 2021.
- Population density-based allocation: The same number of tests are distributed homogeneously across the city, so that each location has the same test rate. Locations with a larger population receive more tests than less densely populated boroughs or MODZCTAs.
- Random allocation: The available tests are distributed randomly across locations. This effectively results in higher per-capita test rates in less densely populated areas.

#### Test Volume

We varied the quantity of available tests from 5% to 400% of the NYC average of 7,184 tests per day (87 tests per 100,000 persons). Changing the quantity of available tests may be understood as either increased capacity or reduced reporting, for example due to a shift toward home testing. From February 2020 to September 2021, an estimated 75 percent of COVID-19 infections went unreported (21). With the emergence of the Omicron sub-variants BA.2.12 and BA.2.12.1, underreporting may be as high as 95 percent (22).

To further understand the role of the geographic test distribution, we also considered “focused testing scenarios”, where individual locations are over-sampled. In these scenarios, 20-100% of all tests were sampled from a single location and the remaining tests were distributed across the remaining locations proportional to population size. We then compared how detection outcomes differed when the new variant emerged in the over-sampled location *versus* one of the other locations.

**Supplementary Figure S2.**
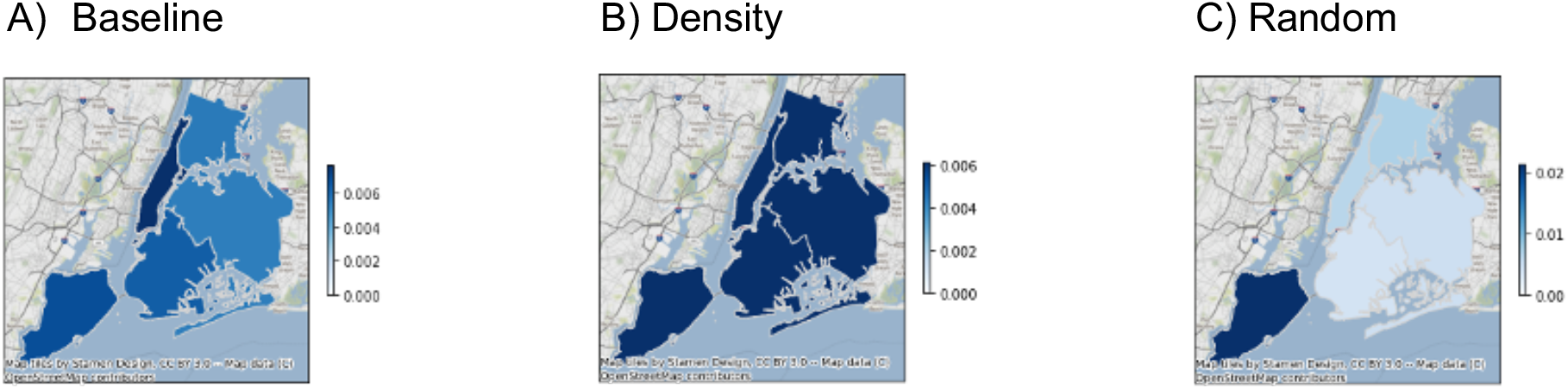
Sample number of tests per capita at the borough level. Boroughs are colored by the proportion of the population that is tested each week under the baseline (A), density-based (B), and random (C) sampling strategy.

#### Sequencing volume

The main analysis considered sequencing probabilities of 5-90% of all positive tests. The sequencing volume for each day of the simulation was therefore determined by the test volume and the number of infections. Raising test volume for a given epidemiologic scenario effectively raises sequencing volume at fixed sequencing probabilities.

In two sensitivity analyses, we sought to isolate the effect of testing and sequencing volumes by placing a cap on sequencing resources:

- Sensitivity analysis 1: At any time point, a fixed number of positive tests was sequenced. If there were fewer positive tests than sequencing resources (due to low prevalence), then all positive tests were sequenced. The maximum sequencing volume was reached when there was a sufficient quantity of positive tests, i.e., sufficiently high prevalence and test volume. The effective sequencing volume did not necessarily vary for different sequencing caps, if prevalence and/or test volume were low.
- Sensitivity analysis 2: A fixed number of tests were assigned as potentials for sequencing, akin to placing a stamp on a subset of test kits. If a test with a stamp was positive, the sample was sequenced. The maximum sequencing volume was achieved when all (stamped) tests were positive, which depended on the number of infections. The effective sequencing volume necessarily varied between the different sequencing caps.

#### Variants

The first variant was modeled to resemble the Delta SARS-CoV-2 variant with an effective transmission probability of 0.2. We then introduced a second variant, which was (i) more transmissible, (ii) had greater and faster immune evasion, or (iii) both. We considered effective transmission probabilities of 0.21-0.5.

#### Context of variant emergence

We simulated the emergence of the second variant at different locations and different times relative to the introduction of the first variant. The first variant was introduced with a single index case in each location, to simulate even spread across the city. The second variant was introduced with one index case in a single location, representing the location of residence of the index case. We simulated all possible introduction locations. Introduction times of the second variant varied from 0 to 150 days after the introduction of the first variant. The introduction times represent different contexts, because of varying prevalence of the first variant and varying numbers of susceptible individuals. We did not distinguish between variants that emerged within the city and those that were imported. The index case of the novel variant in our model could thus represent the first case of a newly emerging variant introduced to NYC from outside the city or a globally undetected variant that either emerged within NYC or was imported from outside before detection.

#### Outcome measures

For each simulation, we computed three primary outcome measures. The *time to detection* was defined as the number of days between the introduction and detection (first sequenced case) of the second variant. *Cumulative undetected infections* measured the total number of people who were exposed to (i.e., infected by) the second variant in NYC by the time the new variant was detected, including individuals who at detection time were in the latent phase prior to infectiousness, infectious, recovered, or susceptible to reinfection. Finally, we computed the *standard deviation of the cumulative undetected infections* across locations as an estimate of the geographic variation in disease burden.

## Supplement 2: Sensitivity analysis of fixed sampling volumes

**Supplementary Figure S3.**
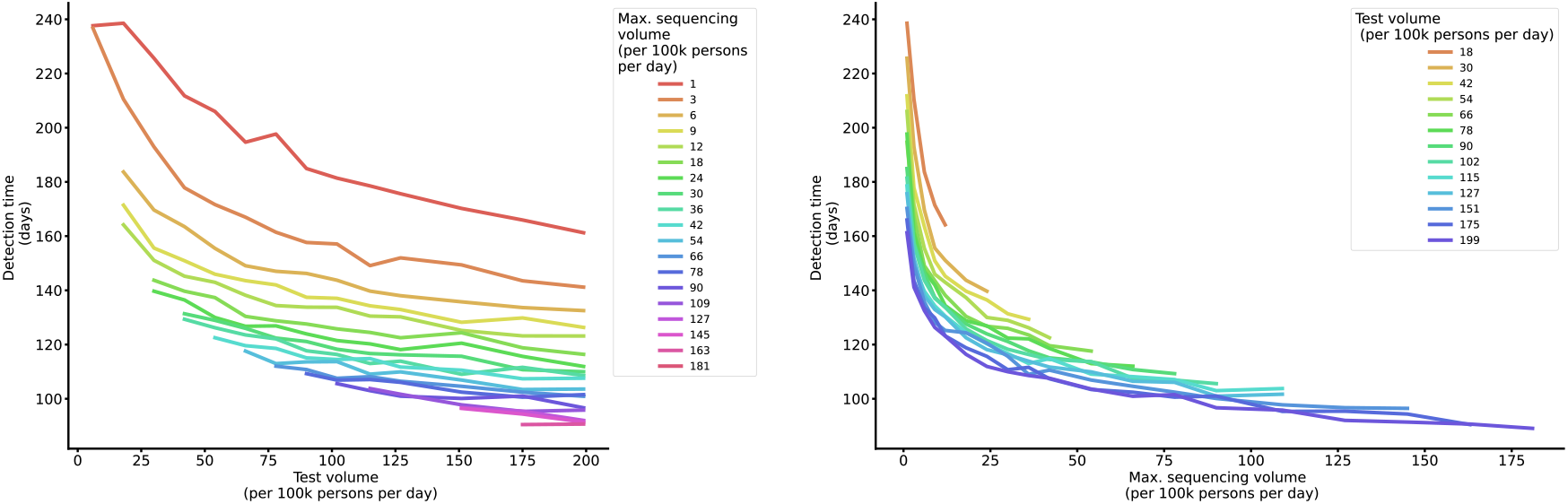
Detection time by fixed volumes of test and sequencing quantities. Lines depict the mean duration between variant introduction and detection in days (A) as a function of daily testing volume, colored by the maximum sequencing volume, and (B) as a function of maximum sequencing volume, colored by the test volume.

## Supplement 3: Role of introduction location

**Supplementary Figure S4.**
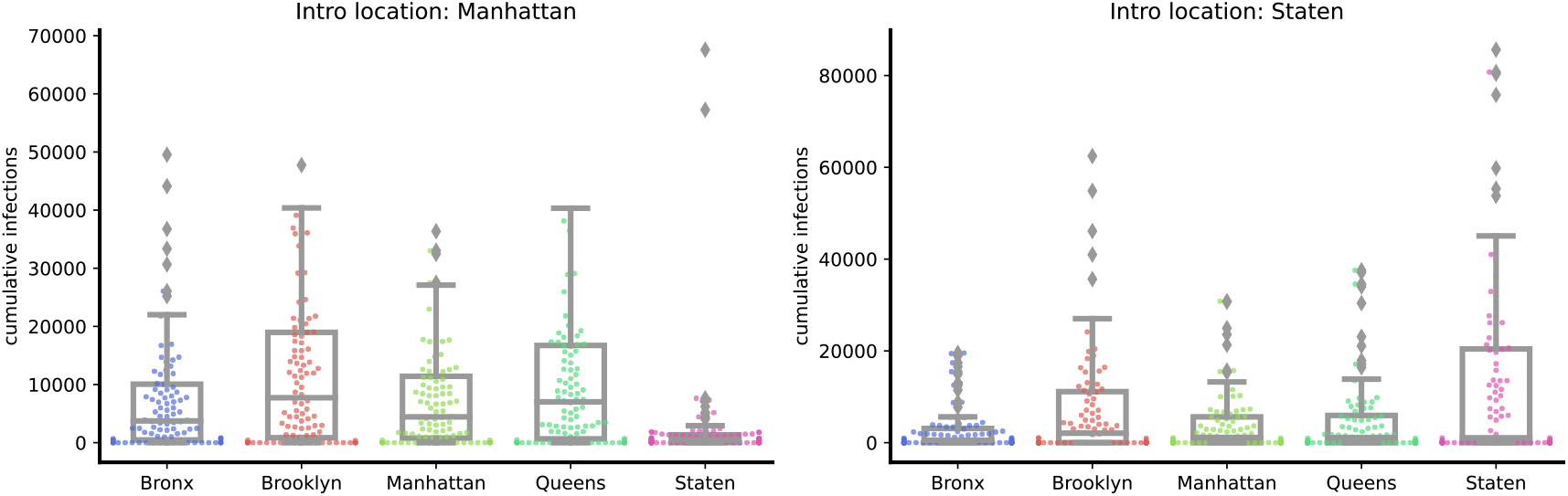
Cumulative infections by borough for introduction locations Manhattan and Staten Island. Points depict the number of cumulative infections in each borough at detection time (at variant introduction 50 days after the prior variant, baseline distribution of tests, 30% of baseline test quantity, and sequencing rate 10%). Boxes and whiskers depict the minimum, lower 25%, median, upper 75%, and maximum cumulative infections.

**Supplementary Figure S5.**
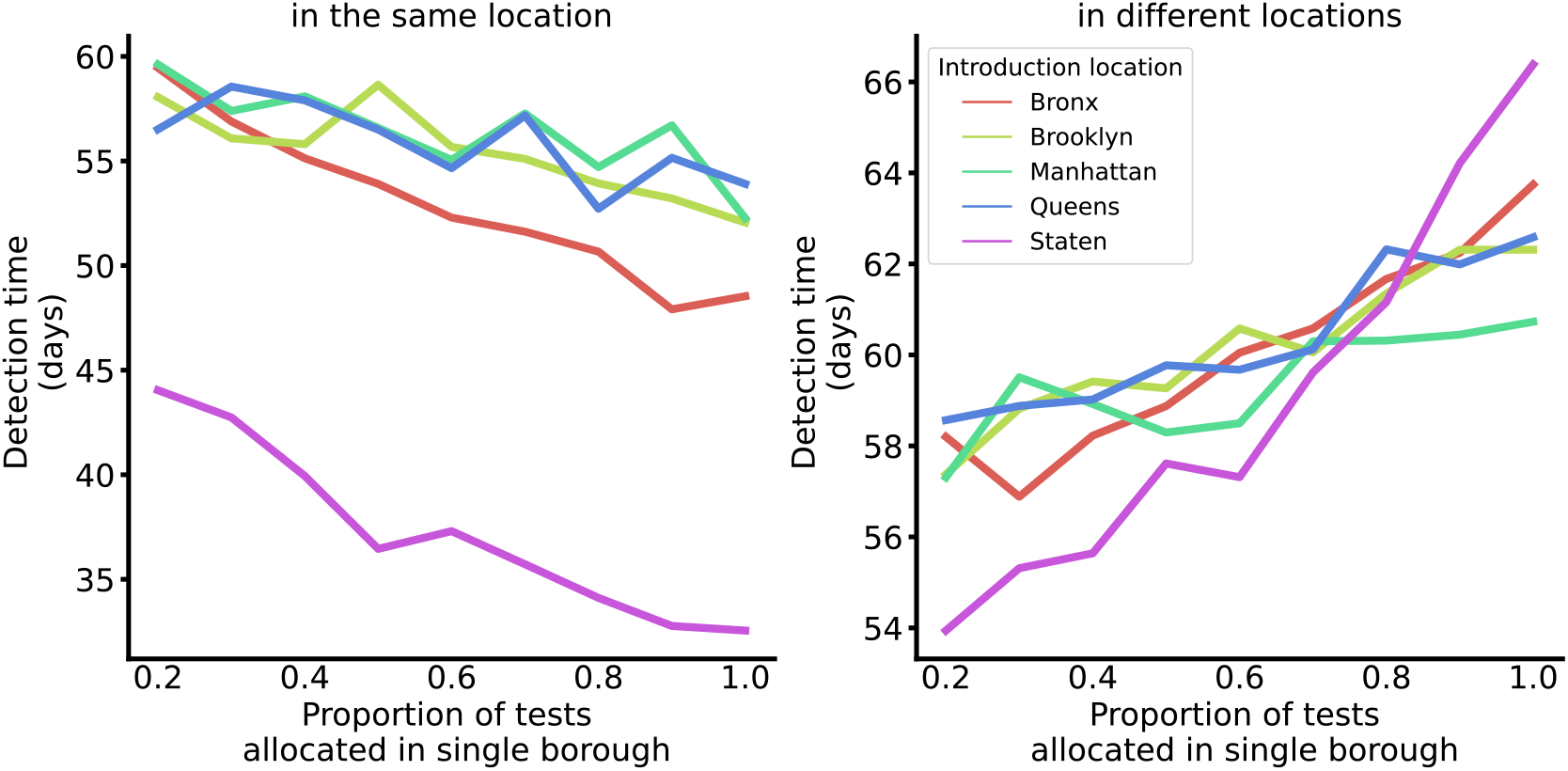
Detection time by proportion of tests allocated in a single location, by introduction location. Lines depict the average detection time for scenarios where between 20% and 100% of tests are sampled from a single location, and the remaining tests are evenly distributed across the remaining locations by population size. The sub-plots distinguish between scenarios where the variant emerged in the primary allocation location, i.e., test over-sampling and emergence occurred in the same location (left), and scenarios where the variant emerged in one of the other locations, i.e., test over-sampling and emergence occurred in different locations (right).

**Supplementary Figure S6.**
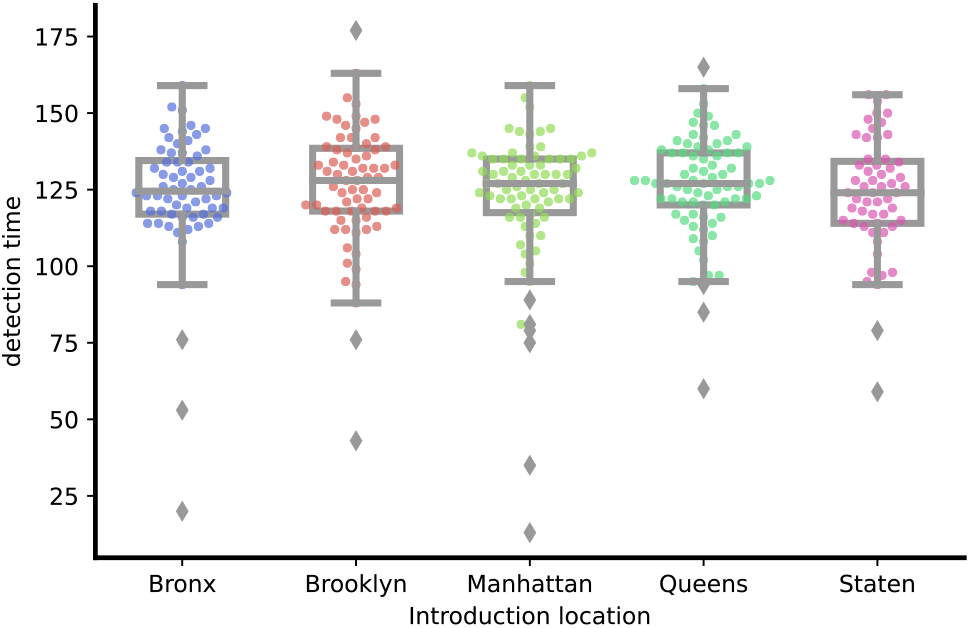
Detection times by introduction location. Points depict the detection time in days for each introduction location (at variant introduction 50 days after the prior variant, baseline distribution of tests, 30% of baseline test quantity, and sequencing rate 10%). Boxes and whiskers depict the minimum, lower 25%, median, upper 75%, and maximum detection times.

## Supplement 4: Social equity

We explored the relationship between socioeconomic variables and the average cumulative undetected exposures at the time of new variant detection across all NYC zip codes (**Supplementary Fig. S7**). We observed a weak negative correlation (Pearson correlation coefficient *-0*.*39*), with cumulative exposures decreasing with increasing household income, possibly due to lower population density in wealthier neighborhoods.

**Supplementary Figure S7.**
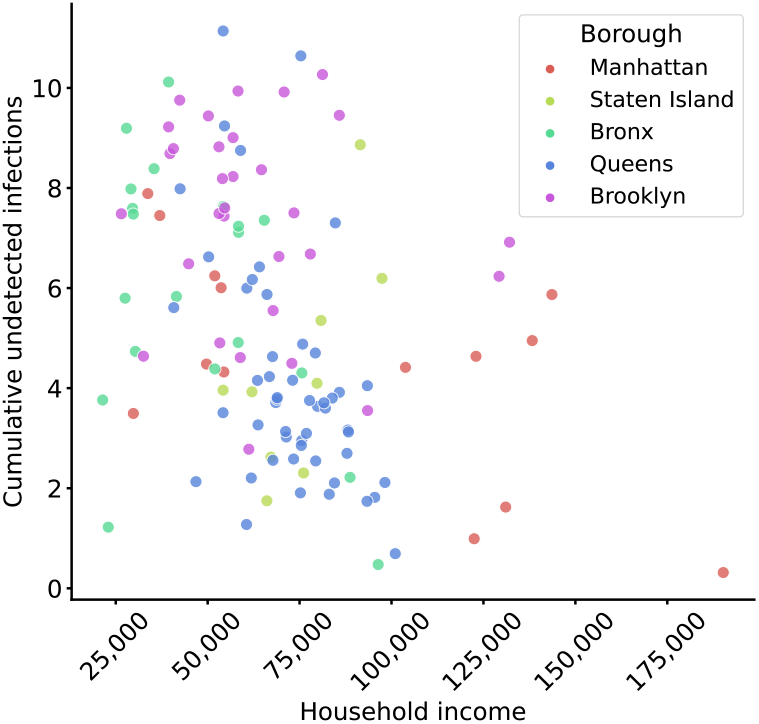
Cumulative undetected exposures by average household income. Points depict the mean cumulative number of infections by household income across 625 simulations (5 simulations for each introduction location) when testing is distributed according to the baseline testing strategy. Colors depict the five boroughs.

## References

1. Walensky RP, Walke HT, Fauci AS. SARS-CoV-2 Variants of Concern in the United States—Challenges and Opportunities. JAMA. 2021 Mar 16;325(11):1037–8.

2. Inzaule SC, Tessema SK, Kebede Y, Ogwell Ouma AE, Nkengasong JN. Genomic-informed pathogen surveillance in Africa: opportunities and challenges. Lancet Infect Dis. 2021 Sep 1;21(9):e281–9.

3. Moderna Announces Omicron-Containing Bivalent Booster Candidate mRNA-1273.214 Demonstrates Superior Antibody Response Against Omicron [Internet]. [cited 2022 Jun 9]. Available from: https://investors.modernatx.com/news/news-details/2022/Moderna-Announces-Omicron-Containing-Bivalent-Booster-Candidate-mRNA-1273.214-Demonstrates-Superior-Antibody-Response-Against-Omicron/default.aspx

4. Viana R, Moyo S, Amoako DG, Tegally H, Scheepers C, Althaus CL, et al. Rapid epidemic expansion of the SARS-CoV-2 Omicron variant in southern Africa. Nature. 2022;603(7902):679–86.

5. Robishaw JD, Alter SM, Solano JJ, Shih RD, DeMets DL, Maki DG, et al. Genomic surveillance to combat COVID-19: challenges and opportunities. Lancet Microbe. 2021 Sep 1;2(9):e481–4.

6. Chen Z, Azman AS, Chen X, Zou J, Tian Y, Sun R, et al. Global landscape of SARS-CoV-2 genomic surveillance and data sharing. Nat Genet. 2022 Apr;54(4):499–507.

7. WHO. Guidance for surveillance of SARS-CoV-2 variants: Interim guidance, 9 August 2021 [Internet]. World Health Organization; 2021 Aug [cited 2022 Nov 19] p. 21. Available from: https://www.who.int/publications-detail-redirect/WHO_2019-nCoV_surveillance_variants

8. ECDC. Guidance for representative and targeted genomic SARS-CoV-2 monitoring. European Centre for Disease Prevention and Control. 2021 May 3;

9. Han AX, Toporowski A, Sacks JA, Perkins MD, Briand S, van Kerkhove M, et al. SARS-CoV-2 diagnostic testing rates determine the sensitivity of genomic surveillance programs. Nat Genet. 2023 Jan;55(1):26–33.

10. Vasylyeva TI, Fang CE, Su M, Havens JL, Parker E, Wang JC, et al. Introduction and Establishment of SARS-CoV-2 Gamma Variant in New York City in Early 2021 [Internet]. medRxiv; 2022 [cited 2022 May 29]. p. 2022.04.15.22273909. Available from: https://www.medrxiv.org/content/10.1101/2022.04.15.22273909v1

11. NYC Coronavirus Disease 2019 (COVID-19) Data [Internet]. NYC Department of Health and Mental Hygiene; 2022 [cited 2022 May 25]. Available from: https://github.com/nychealth/coronavirus-data

12. Maas P. Facebook Disaster Maps: Aggregate Insights for Crisis Response & Recovery. In: Proceedings of the 25th ACM SIGKDD International Conference on Knowledge Discovery & Data Mining [Internet]. New York, NY, USA: Association for Computing Machinery; 2019 [cited 2022 Nov 22]. p. 3173. (KDD ‘19). Available from: https://doi.org/10.1145/3292500.3340412

13. Subissi L, von Gottberg A, Thukral L, Worp N, Oude Munnink BB, Rathore S, et al. An early warning system for emerging SARS-CoV-2 variants. Nat Med. 2022 May 30;

14. Pond EN, Rutkow L, Blauer B, Aliseda Alonso A, Bertran de Lis S, Nuzzo JB. Disparities in SARS-CoV-2 Testing for Hispanic/Latino Populations: An Analysis of State-Published Demographic Data. J Public Health Manag Pract. 2022 Aug;28(4):330.

15. Martin EG. Integrating Health Equity and Efficiency Principles in Distribution Systems: Lessons From Mailing COVID-19 Tests. J Public Health Manag Pract. 2022;28(4):327–9.

16. McPhearson T, Grabowski Z, Herreros-Cantis P, Mustafa A, Ortiz L, Kennedy C, et al. Pandemic Injustice: Spatial and Social Distributions of COVID-19 in the US Epicenter. J Extreme Events. 2020 Dec;07(04):2150007.

17. McDonald SA, Devleesschauwer B, Wallinga J. The impact of individual-level heterogeneity on estimated infectious disease burden: a simulation study. Popul Health Metr. 2016 Dec 1;14:47.

18. Rodriguez-Diaz CE, Guilamo-Ramos V, Mena L, Hall E, Honermann B, Crowley JS, et al. Risk for COVID-19 infection and death among Latinos in the United States: examining heterogeneity in transmission dynamics. Ann Epidemiol. 2020 Dec;52:46-53.e2.

19. Booth A, Reed AB, Ponzo S, Yassaee A, Aral M, Plans D, et al. Population risk factors for severe disease and mortality in COVID-19: A global systematic review and meta-analysis. PloS One. 2021;16(3):e0247461.

20. Wohl S, Lee EC, DiPrete BL, Lessler J. Sample Size Calculations for Variant Surveillance in the Presence of Biological and Systematic Biases. medRxiv. 2022 Jan 1;2021.12.30.21268453.

21. CDC. Cases, Data, and Surveillance [Internet]. Centers for Disease Control and Prevention. 2020 [cited 2022 May 26]. Available from: https://www.cdc.gov/coronavirus/2019-ncov/cases-updates/burden.html

22. Mandavilli A. Two new Omicron subvariants are spreading quickly in New York State. The New York Times [Internet]. 2022 Apr 13 [cited 2022 May 26]; Available from: https://www.nytimes.com/live/2022/04/13/world/covid-19-mandates-cases-vaccine

